# Computer-assisted analysis of routine EEG to identify hidden biomarkers of epilepsy: protocol for a systematic review

**DOI:** 10.1101/2022.06.05.22275999

**Authors:** Émile Lemoine, Joel Neves Briard, Bastien Rioux, Renata Podbielski, Bénédicte Nauche, Denahin Toffa, Mark Keezer, Frédéric Lesage, Dang K. Nguyen, Elie Bou Assi

**Affiliations:** Department of Neurosciences, University of Montreal, Canada; Institute of biomedical engineering, Polytechnique Montreal, Canada; University of Montreal Hospital Center’s Research Center, Canada; School of Public Health, University of Montreal, Canada; Stichting Epilepsie Instellingen Nederland (SEIN), Heemstede, The Netherlands

**Keywords:** Epilepsy, Electroencephalogram, Machine Learning, Diagnosis, Computer-assisted, Biomarker

## Abstract

**Background:** The diagnosis of epilepsy frequently relies on the visual interpretation of the electroencephalogram (EEG) by a neurologist. The hallmark of epilepsy on EEG is the interictal epileptiform discharge (IED). This marker lacks sensitivity: it is only captured in a small percentage of 30-minute routine EEGs in patients with epilepsy. In the past three decades, there has been growing interest in the use of computational methods to analyze the EEG without relying on the detection of IEDs, but none have made it to the clinical practice. We aim to review the diagnostic accuracy of quantitative methods applied to ambulatory EEG analysis to guide the diagnosis and management of epilepsy.

**Methods:** The protocol complies with the recommendations for systematic reviews of diagnostic test accuracy by Cochrane. We will search MEDLINE, EMBASE, EBM reviews, IEEE Explore along with grey literature for articles, conference papers and conference abstracts published after 1961. We will include observational studies that present a computational method to analyze the EEG for the diagnosis of epilepsy in adults or children without relying on the identification of IEDs or seizures. The reference standard is the diagnosis of epilepsy by a physician. We will report the estimated pooled sensitivity and specificity, and receiver operating characteristic area-under-the-curve (ROC AUC) for each marker. If possible, we will perform a meta-analysis of the sensitivity and specificity and ROC AUC for each individual marker. We will assess the risk of bias using an adapted QUADAS-2 tool. We will also describe the algorithms used for signal processing, feature extraction and predictive modeling, and comment on the reproducibility of the different studies.

**Discussion:** Despite the promise to unveil epileptiform patterns that cannot be seen by the naked eye, computational analysis of ambulatory EEG has not yet been successfully translated to the clinical setting. We hope to produce recommendations for future studies on computer-assisted EEG interpretation for the diagnosis and management of epilepsy.

**Systematic review registration:** PROSPERO #292261

## Background

Epilepsy is characterized by an enduring propensity towards epileptic seizures—transient neurological manifestations provoked by a state of abnormal and excessive neuronal activity in the brain^1^. Epilepsy affects over 65 millions of people worldwide, and 10% of the population will experience at least one seizure in their lifetime^2,3^. Epileptic seizures can lead to fractures, road accidents, isolation, anxiety, cognitive decline, and death^4^. In specialized-care settings, the first anti-seizure medication (ASM) achieves seizure freedom in approximately 47% of patients^5^. A prompt diagnosis is key in the prevention of epilepsy-related morbidity and mortality^4^.

A history of epileptic seizures or a high recurrence risk after a single seizure are the basis for the definition of epilepsy by the International League Against Epilepsy (ILAE)^1^. Ancillary tests are often needed to estimate seizure recurrence risk after a single seizure. These include the neurological examination, neuroimaging, and the electroencephalogram (EEG).

An EEG records the electrical activity of the brain. It is recommended that all patients who present with a first unprovoked seizure or with new diagnosis of epilepsy undergo an EEG^6,7^. The initial EEG is generally performed with electrodes applied to the patient’s scalp (scalp EEG or *routine EEG*) for a duration of 20–40 minutes^8^. The EEG tracing is then interpreted visually by a neurologist, who attempts to identify interictal epileptiform discharges (IEDs; *aka* spikes). IEDs are brief (20–200ms) sharp discharges, clearly emerging from background oscillations, often negative in polarity and sometimes followed by a typical slow wave^8^. The presence of interictal spikes on the EEG is considered a hallmark of epilepsy, as it represents a strong predictor of seizure recurrence^9,10^. Furthermore, the identification of interictal spikes can help localize an epileptic focus that may be amenable to surgical resection, and can guide the withdrawal of ASMs in patients after a prolonged period of seizure freedom^11,12^.

The interictal spike has several limitations. It occurs very sporadically: in patients with epilepsy, only 29 – 55% of routine EEGs will capture these transient abnormalities^8^. After a first unprovoked seizure in adults, the sensitivity of a single routine EEG for detecting epileptiform abnormalities is only 17%^9^. Furthermore, their identification is somewhat subjective: the percent agreement between EEG experts is around 76%^13^. Many physiological transient discharges can be misinterpreted as epileptiform spikes. This can lead to the erroneous diagnosis of epilepsy, with sometimes important consequences^14,15^. In patients labelled with drug-resistant epilepsy, over 25% may have had an erroneous diagnosis as a result of both inadequate history taking and misinterpretation of the EEG^16^. Despite the abundant information on brain activity recorded by the EEG, no other interictal anomalies have been validated for use in clinical settings^1,17,18^.

Compared to other neuroimaging modalities, a scalp EEG is inexpensive, easy to acquire, and confers functional information with high temporal resolution^19,20^. Moreover, great effort was put in the last decade by the ILAE in standardizing the equipment, recording and storage of EEG data^10,21^. Decades of research have demonstrated that the automated analysis of EEG can identify hidden differences between with epilepsy and non-epileptic subjects in terms of connectivity^22–24^, signal predictability and complexity^25,26^, spectral power^27,28^, and chaoticity^29^. Computational analysis of EEG holds the promise of extracting information that is invisible to the naked eye of the human interpreter, in an objective and reproducible manner. Discovering new, non-visible markers of epilepsy could increase the diagnostic yield of the EEG, improve its accessibility, and reduce costs, especially in settings where the expertise of a fellowship-trained neurophysiologist is unavailable^18,30^. In spite of this, none of the proposed non-visible markers of epilepsy have made it into clinical practice^10,31^.

We will perform a systematic review of diagnostic test accuracy for automated methods of EEG analysis to distinguish between patients with and without epilepsy without relying on the detection of spikes and seizures. The questions that this review addresses are the following: What is the current evidence on the performances of automatically extracted hidden markers of epilepsy for the diagnosis of epilepsy? And what are the different algorithms that have been tested and how does their diagnostic accuracy compare?

## Methods

### Study design

This will be a systematic review and meta-analysis following guidance from the Cochrane Diagnostic Test Accuracy group. We will report the results according to the PRISMA statement for diagnostic test accuracy (PRISMA-DTA)^32^.

### Study selection criteria

#### Type of studies

We will include all studies that describe a computed marker of epilepsy on routine (scalp) EEG which does not explicitly rely on the identification of interictal spikes or ictal activity (seizures). Studies must compare the EEG signal of individuals with and without epilepsy. We will include retrospective or prospective comparative studies enabling the assessment of diagnostic accuracy (cohort or case-control studies). We will exclude studies reporting data on non-human animals only, studies that include only intracranial or critical care EEG recordings, studies that do not include both individuals with and without epilepsy, and studies that are focused solely on seizure/spike detection or on short-term (<24h) seizure prediction. For studies that include multiple EEG types, we will only extract data that meet the inclusion criteria. We restricted the search to studies published after 1961 (the first use of digital EEG)^33^. There are no restrictions for language.

#### Population

Our population of interest is individuals undergoing routine EEG in a clinical or research setting. A routine EEG is defined as a 20-to 60-minute scalp recording using the international 10–20 electrodes system, with or without prior sleep deprivation. There is no restriction for age groups or diagnoses.

#### Reference standard

We defined the reference standard as the diagnosis of epilepsy by a physician based on criteria specified by the authors (clinical or para-clinical). These criteria must accord with the definition of epilepsy by the ILAE: having had at least one seizure and long-term enduring predisposition to other unprovoked seizures^1,34^.

#### Index test

The index test is a characteristic or feature which is computationally extracted from the EEG signal to identify patients with epilepsy, without relying on detecting IEDs or seizures. These include measures of connectivity, entropy, chaoticity, and power spectrum density^35^. Also included are statistical models that combine several features or models that take as input the raw or processed EEG.

### Search strategy

The search strategy (**Appendix 1**) was developed by two medical librarians specialized in systematic reviews (BN and RP), and peer-reviewed by a senior colleague. We will search MEDLINE (Ovid), EMBASE (Ovid), EBM reviews (Ovid), IEEE Explore along with grey literature for articles, conference papers and conference abstracts. We will use the Covidence platform (Melbourne, Australia) to manage our data for eligibility assessment, selection, and data collection. Two independent reviewers (EL, and either JNB or BR) will screen the records for eligibility using their title and abstract. Any item selected by either reviewer will proceed to the next phase. This process will be repeated on the screened items, this time by consulting the items’ full text. A third, senior reviewer (EBA) will settle conflicts as necessary during the final selection.

### Data items

Data collection will be performed using Covidence by two independent reviewers (EL and JNB/BR), and conflicts will be resolved by a third author (EBA). Authors of the primary study will be contacted if the required data are not available in the original publication. Data collection will include the following information:

1. Title and authors of the study, country of sampling, year of publication;
2. Study type: retrospective vs. prospective, design (cohort, case control);
3. Study sample: exclusion and inclusion criteria, number of screened and included patients;
4. Data collection:
  a. Number of patients, number of EEGs, duration of EEG recordings, use of activation procedures (hyperventilation, photic stimulation, sleep deprivation), setting of recording (hospitalized or ambulatory), whether the same protocol was used for all patients;
  b. Number of electrodes, sampling frequency;
  c. If public dataset: reference to the original dataset, dataset name, exclusion/inclusion criteria used on the EEG segments from the dataset;
  d. Participant characteristics: age, sex, comorbidities, number of ASM, age of first seizure;
5. Reference standard: whether a predefined reference standard was used, definition of reference standard, whether all patients underwent the same reference standard, time lapse between reference standard and EEG;
6. Index test:
  a. Pre-processing: artifact detection and removal (automated or manual), filtering method, filtering frequencies, segmentation protocol (whole EEG vs. EEG segments, window size, overlapping vs. non-overlapping segments, manual vs. automated selection of segments), channel selection;
  b. Feature extraction and selection: multi-channel vs. single channel, number of channels selected, whether feature selection was performed, feature extraction algorithm, feature selection method, whether feature selection was applied to data before vs. after excluding testing data;
  c. Classification: algorithm(s) used for classification, testing methodology (cross-validation vs. held out testing set);
  d. Metric used to report diagnostic performances: ROC AUC, accuracy/sensibility/specificity, F_1_-score, reporting of confidence intervals (CI);
7. Diagnostic performances: number of true positives, number of true negatives, number of false positives, number of false negatives, reported accuracy, reported sensitivity, reported specificity, reported F_1_-score, reported ROC AUC (if more than one index test is performed on the same patient, we will only consider the first test);
8. Reproducibility: whether every data processing step is detailed, whether methods can be reproduced easily, data availability, code availability, open-source computer libraries referenced.

### Risk of bias

The risk of bias of all included studies will be assessed through an adapted version of the QUADAS-2 tool^36^. Risk of bias for each of the following four elements will be evaluated by two independent reviewers (EL and JNB/BR) as low, high, or unclear. Conflicts will be resolved by a third author (EBA). In addition, all publicly available datasets used by at least one of the included studies will be evaluated with the same tool. The following items will be assessed:

1. Patient selection
  a. Is the population representative of clinical practice?
  b. Are inclusion and exclusion criteria identical for cases (patients with epilepsy) and controls?
  c. Are withdrawals explained and appropriate? If individual EEG segments were excluded, were the same criteria used for all segments?
2. Index test
  a. Were the protocols used for recording the EEG identical in all patients, irrespective of the epilepsy diagnosis?
  b. Was the index test validated on an independent sample of patients (patients which were not used to identify the index test’s threshold or train the learning algorithm)?
3. Reference standard
  a. Are the criteria used for the diagnosis of epilepsy specified and acceptable (likely to correctly classify the target condition)?
  b. Was the reference standard assessment independent and blinded to the index test?
4. Flow and timing
  a. Did the whole sample undergo the reference standard?
  b. Did the whole sample undergo the same reference standard?
  c. Was the time lapse between reference standard and EEG acceptable?
  d. Was the same data used in the index method available at the time of the reference standard?
  e. Were all EEGs included in the analysis?

### Data synthesis

We will provide a table summarizing every published study included in the review, comparing the studies’ design, population, reference standard, dataset size, data processing methods, and diagnostic accuracy. We will also provide a table summarizing the risk of bias for all items in the adapted QUADAS-2 tool, comparing 1) every individual article included in the review, and 2) every public dataset that is used in ≥ 2 studies.

We will describe the number of patients, number of EEGs, duration of EEGs, and the EEG-duration-per-patient ratio across all included studies. We will report the pooled proportion of patients with focal vs. generalized epilepsy, adult vs. children, structural vs. non-structural epilepsy, and with specific epilepsy syndromes. For every publicly available dataset identified during the review, we will report the number of studies that used that dataset in their work.

We will summarize the methods used by the different articles during the pipeline’s algorithm (pre-processing, feature extraction, feature selection, and classification algorithm), along with the proportion of studies that used each method.

#### Analyses

We will estimate the specificity and sensitivity for each study, using the Wilson score to compute 95% CI. For studies with varying thresholds, we will estimate the ROC AUC and 95% CI.

If there are sufficient (≥ 5) studies that report the number of true/false positives and true/false negatives, we will estimate the pooled sensitivity and specificity of each individual marker using a hierarchical, bivariate generalized linear mixed model^37^. This allows us to account for the correlation between specificity and sensitivity in a single study. If ≥ 5 studies report these numbers for varying thresholds, we will estimate the pooled ROC curve using the Rutter and Gatsonis HSROC model^38^. All analyses will be implemented with the R statistical language. A *p*-value <0.05 will be considered statistically significant. Given insufficient data for the pooled estimates, we will only describe the diagnostic performances (sensitivity, specificity, ROC AUC) narratively. We will present the results of the analyses with forest plots.

We will quantify heterogeneity using the variances of the logit specificity and sensitivity, as well as the median odds ratio (median OR)^39^. The median OR is a measure of inter-study variance translated on the OR scale. It corresponds to the increase in the odds of being true positive/negative in a patient/control going from a study with lower sensitivity/specificity to a study with higher sensitivity/specificity. For heterogeneity in the ROC plane, we will compute the area of the 95% prediction ellipse^39^. The median OR and the area of the 95% prediction ellipse are easily obtained and interpreted, and take into account the correlation between a single study’s specificity and sensitivity in contrast to univariate methods like Cochrane’s Q and *I*^237,40^. We will perform subgroup analysis for the following variables: epilepsy type (focal, generalized), epilepsy etiology (structural vs. non-structural), age groups (children (< 18 y.o.), adults (≥ 18 y.o.)), epilepsy syndromes, extracted marker, and dataset used. We will assess heterogeneity for all subgroup analyses. We will consider a study as belonging to a particular subgroup if ≥80% of the studied population belongs to that subgroup. Sensitivity analysis will be conducted for the main analyses by excluding studies with overall high/unclear risk of bias.

Some studies use m ultiple markers to classify patients with epilepsy from controls (*e.g*., as input features for a machine learning algorithm). For each marker that is used in ≥ 2 of such studies, we will evaluate the number of studies for which these markers were identified as “important” (selected for the classification task or statistically significant in separating the two classes) and the ratio between the number of studies in which this marker was extracted vs. identified as important.

Reporting bias for sensitivity and specificity will be evaluated by visual inspection of funnel plots.

## Discussion

The interictal EEG is key in the diagnosis of epilepsy, solely based on the visual identification of interictal spikes^41^. Despite years of research on computational biomarkers of epilepsy, only these spikes are currently used in clinical settings^1,17,18^. This review aims to systematically evaluate the diagnostic performances of hidden interictal markers of epilepsy on EEG, describe the data processing pipelines favored by the researchers to classify the EEG for epilepsy diagnosis, and identify the pitfalls that prevent clinical translation of these algorithms.

Algorithms have gained growing interest in medicine for their potential to assist diagnosis and guide clinical decision-making^42^. EEG analysis is well-suited for this application due to the complex nature of the EEG signal. Automated extraction of new epilepsy markers on routine EEG could lead to reduced rate of misdiagnosis, increased availability in areas without access to an expert neurophysiologist, and more efficient clinical trials. Research on automatic analysis of EEG data is thriving, in part assisted by the recent increase in computational capacities^43–50^. However, automatic analysis of EEG is not mentioned in any of the high-quality clinical practice guidelines systematically reviewed by the ILAE^17^.

In recent years, increased computational capacities have allowed the development of powerful algorithms that can learn complex representations such as medical images and EEG signals^43,51,52^. A growing number of algorithms have now been approved by the United States Food and Drug Administration for assisting in the diagnosis of several diseases^53^. Recent systematic reviews have found that most of the studies on automated diagnosis using artificial intelligence have high risk of bias, mostly due to patient selection methodology and absence of validation on external data^54–56^. Systematic reviews on computer-based clinical-decision support systems also highlight the need for more robust patient selection^57–62^. Translation of technology to clinical practice requires strong evidence based on high quality research. This review is important because it will establish the potential of automatic analysis of EEG as a diagnostic tool for epilepsy, and if evidence to support its use is lacking, it will identify the pitfalls that need to be overcome in future research. Also, by systematically describing current practices that are used by research groups, it will serve as a reference for new researchers in the field.

We anticipate that diagnostic accuracy of automatic analysis of EEG for epilepsy will be hard to estimate because of the high heterogeneity between the different dataset used and between the data processing methodology. We also anticipate high risk of bias in many studies, because of the high volume of “proof-of-concept” studies that emphasize computation performances and algorithm development over rigorous diagnostic study methodology. In these cases, we hope to produce recommendations that will assist in bridging the gap between the development of new automated markers and validation in appropriate populations, for ultimate implementation into clinical practice.

## Supporting information

Appendix 1: Search strategy

## Data Availability

All data produced in the present study are available upon reasonable request to the authors

## List of abbreviations

ASM: anti-seizure medication;
CI: confidence interval;
EEG: electroencephalogram;
IED: interictal epileptiform discharge;
ILAE: International League Against Epilepsy;
ROC AUC: receiver operating-characteristic area-under-the-curve.

## Declarations

MRK and DKN report unrestricted educational grants from UCB and Eisai, and research grants for investigator-initiated studies from UCB and Eisai. Émile Lemoine is supported by a scholarship from the Canadian Institute of Health Research. Dang Nguyen is supported by the Canada Research Chairs Program, the Canadian Institutes of Health Research, and Natural Sciences and Engineering Research Council of Canada. Data collected for this study will be available upon reasonable request.

## Authors’ contributions

EL planned the study, drafted the protocol, reviewed the search strategy, and is the guarantor of the review. DT, FL, DKN, and EBA participated in the design of the study. JNB, BR, DT, MRK, FL, DKN, and EBA provided content expertise and critically reviewed the manuscript and the search strategy. BN and RP designed the search strategy. All authors read and approved the final manuscript.

## References

1. Fisher, R. S. et al. ILAE Official Report: A practical clinical definition of epilepsy. Epilepsia 55, 475–482 (2014).

2. Ngugi, A. K., Bottomley, C., Kleinschmidt, I., Sander, J. W. & Newton, C. R. Estimation of the burden of active and life-time epilepsy: a meta-analytic approach. Epilepsia 51, 883–890 (2010).

3. Hauser, W. A. & Beghi, E. First seizure definitions and worldwide incidence and mortality. Epilepsia 49 Suppl 1, 8–12 (2008).

4. Devinsky, O., Spruill, T., Thurman, D. & Friedman, D. Recognizing and preventing epilepsy-related mortality: A call for action. Neurology 86, 779–786 (2016).

5. Kwan, P. & Brodie, M. J. Early identification of refractory epilepsy. N Engl J Med 342, 314–319 (2000).

6. Krumholz, A. et al. Practice Parameter: evaluating an apparent unprovoked first seizure in adults (an evidence-based review): report of the Quality Standards Subcommittee of the American Academy of Neurology and the American Epilepsy Society. Neurology 69, 1996–2007 (2007).

7. Hirtz, D. et al. Practice parameter: evaluating a first nonfebrile seizure in children: report of the quality standards subcommittee of the American Academy of Neurology, The Child Neurology Society, and The American Epilepsy Society. Neurology 55, 616–623 (2000).

8. Pillai, J. & Sperling, M. R. Interictal EEG and the Diagnosis of Epilepsy. Epilepsia 47, 14–22 (2006).

9. Bouma, H. K., Labos, C., Gore, G. C., Wolfson, C. & Keezer, M. R. The diagnostic accuracy of routine electroencephalography after a first unprovoked seizure. European Journal of Neurology 23, 455–463 (2016).

10. Tatum, W. O. et al. Clinical utility of EEG in diagnosing and monitoring epilepsy in adults. Clinical Neurophysiology 129, 1056–1082 (2018).

11. Lamberink, H. J. et al. Individualised prediction model of seizure recurrence and long-term outcomes after withdrawal of antiepileptic drugs in seizure-free patients: a systematic review and individual participant data meta-analysis. Lancet Neurol 16, 523–531 (2017).

12. West, S. et al. Surgery for epilepsy. Cochrane Database of Systematic Reviews (2019) doi:10.1002/14651858.CD010541.pub3.

13. Jing, J. et al. Interrater Reliability of Experts in Identifying Interictal Epileptiform Discharges in Electroencephalograms. JAMA Neurology 77, 49–57 (2020).

14. Amin, U. & Benbadis, S. R. The Role of EEG in the Erroneous Diagnosis of Epilepsy. J Clin Neurophysiol 36, (2019).

15. Kang, J. Y. & Krauss, G. L. Normal Variants Are Commonly Overread as Interictal Epileptiform Abnormalities. J Clin Neurophysiol 36, 257–263 (2019).

16. Smith, D., Defalla, B. A. & Chadwick, D. W. The misdiagnosis of epilepsy and the management of refractory epilepsy in a specialist clinic. QJM 92, 15–23 (1999).

17. Sauro, K. M. et al. The current state of epilepsy guidelines: A systematic review. Epilepsia 57, 13–23 (2016).

18. Engel Jr, J., Bragin, A. & Staba, R. Nonictal EEG biomarkers for diagnosis and treatment. Epilepsia Open 3, 120–126 (2018).

19. DellaBadia Jr, J., Bell, W. L., Keyes Jr, J. W., Mathews, V. P. & Glazier, S. S. Assessment and cost comparison of sleep-deprived EEG, MRI and PET in the prediction of surgical treatment for epilepsy. Seizure 11, 303–309 (2002).

20. Abdelhady, S., Shokri, H., Fathy, M. & wahid el din, mona M. Evaluation of the direct costs of epilepsy in a sample of Egyptian patients following up in Ain Shams University Hospital. The Egyptian Journal of Neurology, Psychiatry and Neurosurgery 56, 112 (2020).

21. Velis, D., Plouin, P., Gotman, J., Da Silva, F. L., & members of the ILAE DMC Subcommittee on Neurophysiology. Recommendations Regarding the Requirements and Applications for Long-term Recordings in Epilepsy. Epilepsia 48, 379–384 (2007).

22. Schmidt, H. et al. A computational biomarker of idiopathic generalized epilepsy from resting state EEG. Epilepsia 57, e200–e204 (2016).

23. Lopes, M. A. et al. Revealing epilepsy type using a computational analysis of interictal EEG. Scientific Reports 9, 10169 (2019).

24. Verhoeven, T. et al. Automated diagnosis of temporal lobe epilepsy in the absence of interictal spikes. NeuroImage: Clinical 17, 10–15 (2018).

25. Ouyang, C.-S., Yang, R.-C., Wu, R.-C., Chiang, C.-T. & Lin, L.-C. Determination of Antiepileptic Drugs Withdrawal Through EEG Hjorth Parameter Analysis. Int. J. Neur. Syst. 30, 2050036 (2020).

26. Zhang, J.-H. et al. Personalized prediction model for seizure-free epilepsy with levetiracetam therapy: a retrospective data analysis using support vector machine. Br J Clin Pharmacol 84, 2615–2624 (2018).

27. Oliva, J. T. & Rosa, J. L. G. Differentiation between Normal and Interictal EEG Using Multitaper Spectral Classifiers. in 2018 International Joint Conference on Neural Networks (IJCNN) 1–8 (2018). doi:10.1109/IJCNN.2018.8489503.

28. Pegg, E. J., Taylor, J. R. & Mohanraj, R. Spectral power of interictal EEG in the diagnosis and prognosis of idiopathic generalized epilepsies. Epilepsy & Behavior 112, 107427 (2020).

29. Jacob, J. E., Sreelatha, V. V., Iype, T., Nair, G. K. & Yohannan, D. G. Diagnosis of epilepsy from interictal EEGs based on chaotic and wavelet transformation. Analog Integrated Circuits and Signal Processing 89, 131–138 (2016).

30. Wahl, B., Cossy-Gantner, A., Germann, S. & Schwalbe, N. R. Artificial intelligence (AI) and global health: how can AI contribute to health in resource-poor settings? BMJ Global Health 3, e000798 (2018).

31. Pitkänen, A. et al. Advances in the development of biomarkers for epilepsy. The Lancet Neurology 15, 843–856 (2016).

32. McInnes, M. D. F. et al. Preferred Reporting Items for a Systematic Review and Meta-analysis of Diagnostic Test Accuracy Studies: The PRISMA-DTA Statement. JAMA 319, 388–396 (2018).

33. November, J. Biomedical computing: Digitizing life in the United States. Biomedical Computing: Digitizing Life in the United States 1–344 (2012).

34. Fisher, R. S. et al. Epileptic seizures and epilepsy: definitions proposed by the International League Against Epilepsy (ILAE) and the International Bureau for Epilepsy (IBE). Epilepsia 46, 470–472 (2005).

35. Supriya, S., Siuly, S., Wang, H. & Zhang, Y. Automated epilepsy detection techniques from electroencephalogram signals: a review study. Health Information Science and Systems 8, 33 (2020).

36. Whiting, P. F. et al. QUADAS-2: a revised tool for the quality assessment of diagnostic accuracy studies. Ann Intern Med 155, 529–536 (2011).

37. Reitsma, J. B. et al. Bivariate analysis of sensitivity and specificity produces informative summary measures in diagnostic reviews. Journal of Clinical Epidemiology 58, 982–990 (2005).

38. Rutter, C. M. & Gatsonis, C. A. A hierarchical regression approach to meta-analysis of diagnostic test accuracy evaluations. Stat Med 20, 2865–2884 (2001).

39. Plana, M. N., Pérez, T. & Zamora, J. New measures improved the reporting of heterogeneity in diagnostic test accuracy reviews: a metaepidemiological study. Journal of Clinical Epidemiology 131, 101–112 (2021).

40. Rücker, G., Schwarzer, G., Carpenter, J. R. & Schumacher, M. Undue reliance on I2 in assessing heterogeneity may mislead. BMC Medical Research Methodology 8, 79 (2008).

41. Smith, S. J. M. EEG in the diagnosis, classification, and management of patients with epilepsy. J Neurol Neurosurg Psychiatry 76, ii2–ii7 (2005).

42. Obermeyer, Z. & Emanuel, E. J. Predicting the Future - Big Data, Machine Learning, and Clinical Medicine. N Engl J Med 375, 1216–1219 (2016).

43. Roy, Y. et al. Deep learning-based electroencephalography analysis: a systematic review. Journal of Neural Engineering 16, 051001 (2019).

44. Craik, A., He, Y. & Contreras-Vidal, J. L. Deep learning for electroencephalogram (EEG) classification tasks: a review. J. Neural Eng. 16, 031001 (2019).

45. Rasheed, K. et al. Machine Learning for Predicting Epileptic Seizures Using EEG Signals: A Review. IEEE Rev. Biomed. Eng. 14, 139–155 (2020).

46. Gemein, L. A. W. et al. Machine-learning-based diagnostics of EEG pathology. NeuroImage 220, 117021 (2020).

47. Mesraoua, B. et al. Electroencephalography in epilepsy: look for what could be beyond the visual inspection. Neurological Sciences 40, 2287–2291 (2019).

48. van Diessen, E. et al. Brain Network Organization in Focal Epilepsy: A Systematic Review and Meta-Analysis. PLOS ONE 9, e114606 (2014).

49. Faiman, I., Smith, S., Hodsoll, J., Young, A. H. & Shotbolt, P. Resting-state EEG for the diagnosis of idiopathic epilepsy and psychogenic nonepileptic seizures: A systematic review. Epilepsy & Behavior 121, 108047 (2021).

50. Pegg, E. J., Taylor, J. R., Keller, S. S. & Mohanraj, R. Interictal structural and functional connectivity in idiopathic generalized epilepsy: A systematic review of graph theoretical studies. Epilepsy & Behavior 106, (2020).

51. Esteva, A. et al. A guide to deep learning in healthcare. Nature Medicine 25, 24–29 (2019).

52. Litjens, G. et al. A survey on deep learning in medical image analysis. Medical Image Analysis 42, 60–88 (2017).

53. FDA Cleared AI Algorithms. American College of Radiology Data Science Institutehttps://models.acrdsi.org.

54. Aggarwal, R. et al. Diagnostic accuracy of deep learning in medical imaging: a systematic review and meta-analysis. npj Digital Medicine 4, 65 (2021).

55. Liu, X. et al. A comparison of deep learning performance against health-care professionals in detecting diseases from medical imaging: a systematic review and meta-analysis. The Lancet Digital Health 1, e271–e297 (2019).

56. Nagendran, M. et al. Artificial intelligence versus clinicians: systematic review of design, reporting standards, and claims of deep learning studies. BMJ vol. 368 m689 (2020).

57. Riches, N. et al. The Effectiveness of Electronic Differential Diagnoses (DDX) Generators: A Systematic Review and Meta-Analysis. PLoS One 11, e0148991 (2016).

58. Bright, T. J. et al. Effect of clinical decision-support systems: a systematic review. Ann Intern Med 157, 29–43 (2012).

59. Jaspers, M. W. M., Smeulers, M., Vermeulen, H. & Peute, L. W. Effects of clinical decision-support systems on practitioner performance and patient outcomes: a synthesis of high-quality systematic review findings. J Am Med Inform Assoc 18, 327–334 (2011).

60. Garg, A. X. et al. Effects of computerized clinical decision support systems on practitioner performance and patient outcomes: a systematic review. JAMA 293, 1223–1238 (2005).

61. Varghese, J., Kleine, M., Gessner, S. I., Sandmann, S. & Dugas, M. Effects of computerized decision support system implementations on patient outcomes in inpatient care: a systematic review. J Am Med Inform Assoc 25, 593–602 (2018).

62. Vasey, B. et al. Association of Clinician Diagnostic Performance With Machine Learning–Based Decision Support Systems: A Systematic Review. JAMA Network Open 4, e211276–e211276 (2021).

