## Appendix 1: Search strategy for "Computer-assisted analysis of routine EEG to identify hidden biomarkers of epilepsy: protocol for a systematic review"

**Medline [OVID]**

Ovid MEDLINE(R) and Epub Ahead of Print, In-Process, In-Data-Review & Other Non-Indexed Citations, Daily and Versions(R) <1946 to December 13, 2021>

| # | Searches | Results |
|---|---|---|
| 1 | exp Electroencephalography/ | 173584 |
| 2 | (EEG* or Electroencephalograph* or "electr* encephalograph*" or "brain wave*").tw,kf. | 111352 |
| 3 | 1 or 2 | 201652 |
| 4 | exp Epilepsy/ | 118716 |
| 5 | Epilep*.tw,kf. | 152323 |
| 6 | (seizure* or convulsion* or infantile spasm*).tw,kf. | 147989 |
| 7 | (BCECTS or BECTS).tw,kf. | 346 |
| 8 | (panayiotopoulos adj2 syndrome*).tw,kf. | 166 |
| 9 | ((Nodding or dravet or doose or may white or fukhura) adj2 (disease* or syndrome*)).tw,kf. | 1407 |
| 10 | (myoencephalopathy ragged red fiber* disease* or MERRF).tw,kf. | 530 |
| 11 | ((Lafora or Unverricht or Landau-Kleffner or Lennox Gastaut) adj2 (disease* or syndrome* or disorder* or seizure*)).tw,kf. | 2534 |
| 12 | or/4-11 | 244612 |
| 13 | exp Algorithms/ | 375058 |
| 14 | Machine learning.tw,kf. | 54804 |
| 15 | ((Deep or hierarchical) adj1 learning).tw,kf. | 25347 |
| 16 | ((transfer* or representation* or network*) adj2 learning).tw,kf. | 7945 |
| 17 | ((artificial or machine or computer or computational) adj2 intelligence).tw,kf. | 19275 |
| 18 | algorithm*.tw,kf. | 299232 |
| 19 | ((data or binary or multiclass or multilabel) adj2 classification).tw,kf. | 4758 |

| | | |
|---|---|---|
| 20 | ((artificial or computational or computer* or convolutional or connectionist or mathematical) adj2 neur* network*).tw,kf. | 28375 |
| 21 | exp Pattern Recognition, Automated/ | 26085 |
| 22 | (Automat* adj2 pattern* adj2 recognition*).tw,kf. | 155 |
| 23 | (Back* propagation* or backpropagation*).tw,kf. | 4397 |
| 24 | exp Bayes Theorem/ | 40554 |
| 25 | (Bayes* adj2 (theorem or learning or analysis or approach* or forecast* or method* or prediction*)).tw,kf. | 21469 |
| 26 | (feature* adj2 (detecti* or extracti* or learning* or ranking* or selection*)).tw,kf. | 21577 |
| 27 | (Fuzzy or neurofuzzy).tw,kf. | 13240 |
| 28 | exp Markov chains/ | 15485 |
| 29 | (Markov adj2 (model* or chain$1 or process*)).tw,kf. | 21918 |
| 30 | K nearest neighbor*.tw,kf. | 3529 |
| 31 | (Kernel$1 adj2 (method* or algorithm* or approach or correlation or estim* or regression or model* or string or tree)).tw,kf. | 3950 |
| 32 | exp Knowledge discovery/ | 130 |
| 33 | (Knowledge adj2 discover*).tw,kf. | 1589 |
| 34 | exp Multifactor Dimensionality Reduction/ | 226 |
| 35 | Dimensionality reduction*.tw,kf. | 3836 |
| 36 | (predicti* adj2 model*).tw,kf. | 79862 |
| 37 | connectom*.tw,kf. | 4980 |
| 38 | neur* decod*.tw,kf. | 361 |
| 39 | (outlier* adj2 detection*).tw,kf. | 893 |
| 40 | Neural networks, computer/ | 35265 |
| 41 | (neural adj2 network*).tw,kf. | 70371 |
| 42 | perceptron*.tw,kf. | 3390 |
| 43 | radial basis function*.tw,kf. | 2359 |
| 44 | random forest*.tw,kf. | 13717 |

| | | |
|---|---|---|
| 45 | recursive feature* elimination*.tw,kf. | 688 |
| 46 | recursive partition*.tw,kf. | 2380 |
| 47 | exp Support Vector Machine/ | 8553 |
| 48 | (vector* adj2 (machine* or classifi* or network* or regression)).tw,kf. | 22248 |
| 49 | support vector*.tw,kf. | 21483 |
| 50 | rough set*.tw,kf. | 397 |
| 51 | ((automat* or electron* or comput* or information or analytic*) adj2 (processing or reasoning)).tw,kf. | 38719 |
| 52 | (quantitative adj2 analys*).tw,kf. | 90324 |
| 53 | (Peak* adj2 (alpha* or frequenc*)).tw,kf. | 5453 |
| 54 | Entrop*.tw,kf. | 45494 |
| 55 | Lyapunov exponent*.tw,kf. | 2179 |
| 56 | Hjorth*.tw,kf. | 184 |
| 57 | Sub-band energ*.tw,kf. | 18 |
| 58 | exp fourier Analysis/ | 17272 |
| 59 | (Fourier* or (cyclic adj2 (analys* or series or transform* or approach*)) or FFT).tw,kf. | 87439 |
| 60 | (Hilbert* adj2 transform*).tw,kf. | 1008 |
| 61 | (dimension* adj2 (fractal* or correlation*)).tw,kf. | 8106 |
| 62 | (Hurst adj2 exponent*).tw,kf. | 575 |
| 63 | exp wavelet analysis/ | 2541 |
| 64 | (Wavelet* adj2 (analysis or processing or transform*)).tw,kf. | 7248 |
| 65 | phase locking value*.tw,kf. | 311 |
| 66 | Fisher information*.tw,kf. | 870 |
| 67 | Dynamic network*.tw,kf. | 1839 |
| 68 | Principal component* analys*.tw,kf. | 47819 |
| 69 | Independant component* analys*.tw,kf. | 2 |
| 70 | Functional connectivit*.tw,kf. | 22171 |

| | | |
|---|---|---|
| 71 | (gradient* boost* or Adaboost*).tw,kf. | 3337 |
| 72 | (QEEG or Quantitative Electroencephalogra*).tw,kf. | 1750 |
| 73 | (chaotic feature* or chaos).tw,kf. | 9755 |
| 74 | comput*.tw,kf. | 958508 |
| 75 | quantitative.tw,kf. | 689806 |
| 76 | or/13-75 | 2378446 |
| 77 | (sensitiv* or diagnos* or predict*).mp. or scor*.tw. or observ*.mp. | 11325259 |
| 78 | di.fs. | 2760821 |
| 79 | or/77-78 | 11325259 |
| 80 | 3 and 12 and 76 and 79 | 5990 |
| 81 | (Animals/ or Models, animal/ or Disease models, animal/) not Humans/ | 4900078 |
| 82 | ((animal or animals or canine* or cat or cats or dog or dogs or feline or hamster* or lamb or lambs or mice or monkey or monkeys or mouse or murine or pig or pigs or piglet* or porcine or primate* or rabbit* or rats or rat or rodent* or sheep* or veterinar*) not (human* or patient* or women or men)).tw,kf. | 3315730 |
| 83 | 81 or 82 | 5542727 |
| 84 | 80 not 83 | 5627 |
| 85 | limit 84 to yr="1961 -Current" | 5627 |

### EMBASE [OVID]

Embase <1974 to 2021 December 13>

| # | Searches | Results |
|---|---|---|
| 1 | exp electroencephalography/ | 124495 |
| 2 | (EEG* or Electroencephalograph* or "electr* encephalograph*" or "brain wave*").tw,kf. | 146325 |
| 3 | 1 or 2 | 206929 |
| 4 | exp epilepsy/ | 251058 |
| 5 | Epilep*.tw,kf. | 214171 |
| 6 | (seizure* or convulsion* or infantile spasm*).tw,kf. | 216888 |
| 7 | (BCECTS or BECTS).tw,kf. | 509 |

| | | |
|---|---|---|
| 8 | (panayiotopoulos adj2 syndrome*).tw,kf. | 249 |
| 9 | ((Nodding or dravet or doose or may white or fukhura) adj2 (disease* or syndrome*)).tw,kf. | 2324 |
| 10 | (myoencephalopathy ragged red fiber* disease* or MERRF).tw,kf. | 711 |
| 11 | ((Lafora or Unverricht or Landau-Kleffner or Lennox Gastaut) adj2 (disease* or syndrome* or disorder* or seizure*)).tw,kf. | 3984 |
| 12 | or/4-11 | 371364 |
| 13 | Machine learning/ | 49774 |
| 14 | Machine learning.tw,kf. | 63858 |
| 15 | ((Deep or hierarchical) adj1 learning).tw,kf. | 28566 |
| 16 | exp network learning/ | 886 |
| 17 | ((transfer* or representation* or network*) adj2 learning).tw,kf. | 8790 |
| 18 | exp artificial intelligence/ | 55153 |
| 19 | ((artificial or machine or computer or computational) adj2 intelligence).tw,kf. | 23056 |
| 20 | exp algorithm/ | 465121 |
| 21 | algorithm*.tw,kf. | 381089 |
| 22 | ((data or binary or multiclass or multilabel) adj2 classification).tw,kf. | 6087 |
| 23 | exp artificial neural network/ | 62826 |
| 24 | ((artificial or computational or computer* or convolutional or connectionist or mathematical) adj2 neur* network*).tw,kf. | 33889 |
| 25 | exp pattern recognition/ or exp automated pattern recognition/ | 68427 |
| 26 | (Automat* adj2 pattern* adj2 recognition*).tw,kf. | 199 |
| 27 | exp back propagation/ | 2553 |
| 28 | (Back* propagation* or backpropagation*).tw,kf. | 5107 |
| 29 | exp Bayesian learning/ | 4303 |
| 30 | (Bayes* adj2 (theorem or learning or analysis or approach* or forecast* or method* or prediction*)).tw,kf. | 24116 |
| 31 | exp Feature detection/ or exp feature extraction/ or exp feature learning/ or exp feature ranking/ or exp feature selection/ | 31030 |

| | | |
|---|---|---|
| 32 | ((feature* or representation) adj2 (detecti* or extracti* or learning* or ranking* or selection*)).tw,kf. | 28097 |
| 33 | exp fuzzy system/ | 4077 |
| 34 | (fuzzy or neurofuzzy).tw,kf. | 16138 |
| 35 | exp Markov chain/ or exp Markov state model/ | 12093 |
| 36 | (Markov adj2 (model* or chain$1 or process*)).tw,kf. | 29000 |
| 37 | exp k nearest neighbor/ | 4553 |
| 38 | K nearest neighbor*.tw,kf. | 4260 |
| 39 | kernel method/ | 6720 |
| 40 | (Kernel$1 adj2 (method* or algorithm* or approach or correlation or estim* or regression or model* or string or tree)).tw,kf. | 4389 |
| 41 | exp Knowledge discovery/ | 727 |
| 42 | (Knowledge adj2 discover*).tw,kf. | 1804 |
| 43 | exp multifactor dimensionality reduction/ | 864 |
| 44 | Dimension* reduction*.tw,kf. | 7086 |
| 45 | (predicti* adj2 model*).tw,kf. | 105404 |
| 46 | connectom*.tw,kf. | 6225 |
| 47 | neur* decod*.tw,kf. | 433 |
| 48 | exp Outlier detection/ | 470 |
| 49 | (outlier* adj2 detection*).tw,kf. | 1010 |
| 50 | exp artificial neural network/ | 62826 |
| 51 | exp Perceptron/ | 2478 |
| 52 | perceptron*.tw,kf. | 3962 |
| 53 | (neural adj2 network*).tw,kf. | 84786 |
| 54 | exp radial basis function/ | 942 |
| 55 | radial bas* function*.tw,kf. | 2927 |
| 56 | exp random forest/ | 14358 |
| 57 | (random adj2 forest*).tw,kf. | 17752 |

| | | |
|---|---|---|
| 58 | exp recursive feature elimination/ | 393 |
| 59 | recursive feature* elimination*.tw,kf. | 860 |
| 60 | exp recursive partitioning/ | 462 |
| 61 | recursive partition*.tw,kf. | 3567 |
| 62 | exp relevance vector machine/ or exp support vector machine/ | 28522 |
| 63 | (vector* adj2 (machine* or classifi* or network* or regression)).tw,kf. | 27021 |
| 64 | support vector*.tw,kf. | 26266 |
| 65 | exp rough set/ | 248 |
| 66 | rough set*.tw,kf. | 531 |
| 67 | exp online analytical processing/ | 187 |
| 68 | ((automat* or electron* or comput* or information or analytic*) adj2 (processing or reasoning)).tw,kf. | 44254 |
| 69 | Quantitative analysis/ | 367570 |
| 70 | (quantitative adj2 analys*).tw,kf. | 113093 |
| 71 | (Peak* adj2 (alpha* or frequenc*)).tw,kf. | 6315 |
| 72 | Entrop*.tw,kf. | 43483 |
| 73 | Lyapunov exponent*.tw,kf. | 1600 |
| 74 | Hjorth*.tw,kf. | 264 |
| 75 | Sub-band energ*.tw,kf. | 23 |
| 76 | exp Fourier analysis/ | 10056 |
| 77 | (Fourier* or (cyclic adj2 (analys* or series or transform* or approach*)) or FFT).tw,kf. | 89584 |
| 78 | Hilbert transform/ | 183 |
| 79 | (Hilbert* adj2 transform*).tw,kf. | 1253 |
| 80 | (dimension* adj2 (fractal* or correlation*)).tw,kf. | 8947 |
| 81 | (Hurst adj2 exponent*).tw,kf. | 555 |
| 82 | exp wavelet transform/ | 2217 |
| 83 | (Wavelet* adj2 (analysis or processing or transform*)).tw,kf. | 9182 |

| | | |
|---|---|---|
| 84 | phase locking value*.tw,kf. | 425 |
| 85 | Fisher information*.tw,kf. | 746 |
| 86 | Dynamic network*.tw,kf. | 1972 |
| 87 | Principal component* analys*.tw,kf. | 58526 |
| 88 | Independent component* analys*.tw,kf. | 7493 |
| 89 | Functional connectivity/ | 21903 |
| 90 | Functional connectivit*.tw,kf. | 30389 |
| 91 | (gradient* boost* or Adaboost*).tw,kf. | 4097 |
| 92 | (QEEG or Quantitative Electroencephalogra*).tw,kf. | 2861 |
| 93 | (chaotic feature* or chaos).tw,kf. | 8412 |
| 94 | comput*.tw,kf. | 1156500 |
| 95 | quantitative.tw,kf. | 852081 |
| 96 | or/13-95 | 2994032 |
| 97 | (sensitiv* or diagnos* or predict*).mp. or scor*.tw. or observ*.mp. | 14413096 |
| 98 | di.fs. | 3343316 |
| 99 | or/97-98 | 14413096 |
| 100 | 3 and 12 and 96 and 99 | 8362 |
| 101 | (exp animal/ or animal experiment/ or nonhuman/) not (exp human/ or human experiment/) | 6801969 |
| 102 | (animal or animals or canine* or dog or dogs or feline or hamster* or lamb or lambs or mice or monkey ormonkeys or mouse or murine or pig or pigs or piglet* or porcine or primate* or rabbit* or rats or rat or rodent* or sheep* or veterinar*).ti,kw,dq,jx. not (human* or patient*).mp. | 2062187 |
| 103 | 101 or 102 | 6872024 |
| 104 | 100 not 103 | 7906 |
| 105 | limit 104 to yr="1961 -Current" | 7890 |
| 106 | limit 105 to embase | 5134 |

### EBM Reviews [OVID]

All EBM Reviews - Cochrane DSR, ACP Journal Club, DARE, CCA, CCTR, CMR, HTA, and NHSEED <executed on December 14>

| # | Searches | Results |
|---|---|---|
| 1 | (EEG* or Electroencephalograph* or "electr* encephalograph*" or "brain wave*").tw,kw,sh. | 12245 |
| 2 | Epilep*.tw,kw,sh. | 10099 |
| 3 | (seizure* or convulsion* or infantile spasm*).tw,kw,sh. | 11675 |
| 4 | (BCECTS or BECTS).tw,kw,sh. | 31 |
| 5 | (panayiotopoulos adj2 syndrome*).tw,kw,sh. | 5 |
| 6 | ((Nodding or dravet or doose or may white or fukhura) adj2 (disease* or syndrome*)).tw,kw,sh. | 413 |
| 7 | (myoencephalopathy ragged red fiber* disease* or MERRF).tw,kw,sh. | 5 |
| 8 | ((Lafora or Unverricht or Landau-Kleffner or Lennox Gastaut) adj2 (disease* or syndrome* or disorder* or seizure*)).tw,kw,sh. | 339 |
| 9 | or/2-8 | 16595 |
| 10 | algorithm*.tw,kw. | 16401 |
| 11 | Machine learning.tw,kw,sh. | 1918 |
| 12 | ((Deep or hierarchical) adj1 learning).tw,kw,sh. | 708 |
| 13 | ((transfer* or representation* or network*) adj2 learning).tw,kw,sh. | 691 |
| 14 | ((artificial or machine or computer or computational) adj2 intelligence).tw,kw,sh. | 827 |
| 15 | algorithm*.tw,kw,sh. | 18549 |
| 16 | ((data or binary or multiclass or multilabel) adj2 classification).tw,kw,sh. | 335 |
| 17 | ((artificial or computational or computer* or connectionist or convolutional or mathematical) adj2 neur* network*).tw,kw,sh. | 782 |
| 18 | (Automat* adj2 pattern* adj2 recognition*).tw,kw,sh. | 15 |
| 19 | (Back* propagation* or backpropagation*).tw,kw,sh. | 66 |
| 20 | (Bayes* adj2 (theorem or learning or analysis or approach* or forecast* or method* or prediction*)).tw,kw,sh. | 1841 |
| 21 | (feature* adj2 (detecti* or extracti* or learning* or ranking* or selection*)).tw,kw,sh. | 607 |

| | | |
|---|---|---|
| 22 | (fuzzy or neurofuzzy).tw,kw,sh. | 197 |
| 23 | (Markov adj2 (model* or chain$1 or process*)).tw,kw,sh. | 4373 |
| 24 | K nearest neighbor*.tw,kw,sh. | 73 |
| 25 | (Kernel$1 adj2 (method* or algorithm* or approach or correlation or estim* or regression or model* or string or tree)).tw,kw,sh. | 90 |
| 26 | (Knowledge adj2 discover*).tw,kw,sh. | 26 |
| 27 | Dimensionality reduction*.tw,kw,sh. | 73 |
| 28 | (predicti* adj2 model*).tw,kw,sh. | 5378 |
| 29 | connectom*.tw,kw,sh. | 308 |
| 30 | neur* decod*.tw,kw,sh. | 2 |
| 31 | (outlier* adj2 detection*).tw,kw,sh. | 14 |
| 32 | perceptron*.tw,kw,sh. | 76 |
| 33 | (neural adj2 network*).tw,kw,sh. | 1672 |
| 34 | radial basis function*.tw,kw,sh. | 39 |
| 35 | random forest*.tw,kw,sh. | 615 |
| 36 | recursive feature* elimination*.tw,kw,sh. | 30 |
| 37 | recursive partition*.tw,kw,sh. | 282 |
| 38 | (vector* adj2 (machine* or classifi* or network* or regression)).tw,kw,sh. | 555 |
| 39 | support vector*.tw,kw,sh. | 544 |
| 40 | rough set*.tw,kw,sh. | 3 |
| 41 | ((automat* or electron* or comput* or information or analytic*) adj2 (processing or reasoning)).tw,kw,sh. | 7510 |
| 42 | (quantitative adj2 analys*).tw,kw,sh. | 8960 |
| 43 | (Peak* adj2 (alpha* or frequenc*)).tw,kw,sh. | 357 |
| 44 | Entrop*.tw,kw,sh. | 951 |
| 45 | Lyapunov exponent*.tw,kw,sh. | 37 |
| 46 | Hjorth*.tw,kw,sh. | 29 |
| 47 | Sub-band energ*.tw,kw,sh. | 0 |

| | | |
|---|---|---|
| 48 | (Fourier* or (cyclic adj2 (analys* or series or transform* or approach*)) or FFT).tw,kw,sh. | 1043 |
| 49 | (Hilbert* adj2 transform*).tw,kw,sh. | 19 |
| 50 | (dimension* adj2 (fractal* or correlation*)).tw,kw,sh. | 184 |
| 51 | (Hurst adj2 exponent*).tw,kw,sh. | 14 |
| 52 | (Wavelet* adj2 (analysis or processing or transform*)).tw,kw,sh. | 126 |
| 53 | phase locking value*.tw,kw,sh. | 11 |
| 54 | Fisher information*.tw,kw,sh. | 7 |
| 55 | Dynamic network*.tw,kw,sh. | 12 |
| 56 | Principal component* analys*.tw,kw,sh. | 1207 |
| 57 | Independant component* analys*.tw,kw,sh. | 0 |
| 58 | Functional connectivit*.tw,kw,sh. | 2220 |
| 59 | (gradient* boost* or Adaboost*).tw,kw,sh. | 168 |
| 60 | (QEEG or Quantitative Electroencephalogra*).tw,kw,sh. | 448 |
| 61 | (chaotic feature* or chaos).tw,kw,sh. | 141 |
| 62 | comput*.tw,kw,sh. | 80820 |
| 63 | quantitative.tw,kw,sh. | 33706 |
| 64 | or/10-63 | 145496 |
| 65 | (sensitiv* or diagnos* or predict*).mp. or scor*.tw. or observ*.mp. | 810011 |
| 66 | di.tw,kw,sh. | 17162 |
| 67 | 65 or 66 | 811399 |
| 68 | 1 and 9 and 64 and 67 | 350 |
| 69 | ((animal or animals or canine* or cat or cats or dog or dogs or feline or hamster* or lamb or lambs or mice or monkey or monkeys or mouse or murine or pig or pigs or piglet* or porcine or primate* or rabbit* or rats or rat or rodent* or sheep* or veterinar*) not (human* or patient* or women or men)).tw,kw,sh. | 5147 |
| 70 | 68 not 69 | 346 |
| 71 | limit 70 to yr="1961 -Current" [Limit not valid in DARE; records were retained] | 321 |
| 72 | remove duplicates from 71 | 315 |

### IEEE Xplore

<executed on December 14>

| | |
|---|---|
| ((((((((All Metadata:predicted OR All Metadata:prediction OR All Metadata:predictions OR All Metadata:predicting OR All Metadata:predictive OR All Metadata:predictor OR All Metadata:predictors OR All Metadata:predicts OR All Metadata:predictability OR All Metadata:predictable OR All Metadata:predictably OR All Metadata:predictively OR All Metadata:predictiveness))) OR ((All Metadata:sensitivity OR All Metadata:sensitively OR All Metadata:sensitiveness OR All Metadata:sensitive OR All Metadata:sensitivities))) OR ((All Metadata:diagnose OR All Metadata:diagnosis OR All Metadata:diagnosed OR All Metadata:diagnoses OR All Metadata:diagnostic OR All Metadata:diagnosing OR All Metadata:diagnosable OR All Metadata:diagnostics OR All Metadata:diagnoseable OR All Metadata:diagnostical OR All Metadata:diagnostician OR All Metadata:diagnosticians OR All Metadata:diagnostically))) AND ((No Keywords Specified))) AND ((No Keywords Specified))) AND ((Index Terms:EEG ) OR (Index Terms:Electroencephalograph*) OR (Index Terms: "electr* encephalograph*") OR (Index Terms: "brain wave") OR (Index Terms:"brain waves"))) OR ((Document Title:EEG) OR (Document Title:Electroencephalograph*) OR (Document Title:"electr* encephalograph*") OR (Document Title:"brain wave") OR (Document Title:"brain waves"))) AND ((Index Terms:epilep*) OR (Document Title:seizure OR Document Title:seizures OR Document Title:convulsion OR Document Title:convulsions OR Document Title:"infantile spasm" OR Document Title:"infantile spasms")) | 2492 |

### Google Scholar (using Publish or Perish)

<executed on December 21>

| | |
|---|---|
| Electroencephalogram epilepsy [title], machine learning algorithm* diagnos* [keywords] | 32 selected articles out of 32 |
| Electroencephalography epilepsy [title], machine learning algorithm* diagnos* [keywords] | 21 selected article out of 21 |
| EEG epilepsy [title], machine learning algorithm* diagnos* [keywords] | 433 sur 433 |

### Grey literature

#### Alberta: Health evidence reviews

https://www.alberta.ca/health-evidence-reviews.aspx

| | |
|---|---|
| Electroencephalography | 0 selected articles out of 1 |
| EEG | 0 selected articles out of 3 |

#### Canadian Agency for Drug and Technologies in Health

https://www.cadth.ca/search?keywords

| | |
|---|---|
| Electroencephalography | 0 selected articles out of 1 |
| EEG | 0 selected articles out of 4 |

**Health Quality Council of Alberta**

https://hqca.ca/studies-and-reviews/

| | |
|---|---|
| Electroencephalography | 0 selected articles out of 0 |
| EEG | 0 selected articles out of 0 |

**Health Quality Ontario: Health Technology Assessment**

Quality Standards - Health Quality Ontario (HQO) (hqontario.ca)

| | |
|---|---|
| Electroencephalography | 1 selected article out of 7 |
| EEG | 1 selected article out of 5 |

**INESS**

https://www.inesss.qc.ca/en/publications/publications.html?tx_solr%5Bq%5D=EEG

| | |
|---|---|
| électroencéphalographie | 0 selected articles out of 5 |
| EEG | 0 selected articles out of 0 |

**McGill University Health Centre (MUHC). Technology Assessment Unit Reports**

https://muhc.ca/tau/page/tau-reports

| | |
|---|---|
| Electroencephalography | 0 selected article out of 0 |
| EEG | 0 selected articles out of 3 |

**Newfoundland & Labrador Centre For Applied Health Research**

http://www.nlcahr.mun.ca/CHRSP/CompletedCHRSP.php

| | |
|---|---|
| Electroencephalography AND epilepsy | 0 selected articles out of 37 |
| Electroencephalogram AND epilepsy | 0 selected articles out of 34 |
| EEG AND epilepsy | 0 selected articles out of 28 |

**The Ottawa Hospital Research institute: Knowledge Synthesis Group**

http://www.ohri.ca/ksgroup/

| | |
|---|---|
| Electroencephalography | 0 selected articles out of 0 |
| Electroencephalogram | 0 selected articles out of 0 |
| EEG AND epilepsy | 0 selected articles out of 7 |

**Programs for Assessment of Technology in Health**

https://www.path-hta.com/research-1

| | |
|---|---|
| Electroencephalography | 0 selected articles out of 0 |
| Electroencephalogram | 0 selected articles out of 0 |
| EEG | 0 selected articles out of 0 |

**The International Network of Agencies for Health Technology Assessment**

**Publications - INAHTA**

| | |
|---|---|
| Electroencephalography | 0 selected articles out of 1 |
| Electroencephalogram | 0 selected articles out of 4 |
| EEG | 0 selected articles out of 4 |

**Horizon Scanning**

**Horizon Scanning - Australia and New Zealand Horizon Scanning Network - Technologies Assessed**

| | |
|---|---|
| Electroencephalography | 0 selected articles out of 1 |
| Electroencephalogram | 0 selected articles out of 0 |
| EEG | 0 selected articles out of 0 |

**Austrian Academy of Sciences**

https://www.oeaw.ac.at/en/

| | |
|---|---|
| Electroencephalography | 0 selected articles out of 0 |
| Electroencephalogram | 0 selected articles out of 0 |
| EEG | 0 selected articles out of 2 |

**Austrian Institute Of Health Technology Assessment**

Welcome to Repository of AIHTA GmbH - Repository of AIHTA GmbH (lbg.ac.at)

| | |
|---|---|
| Electroencephalography | 0 selected articles out of 4 |
| Electroencephalogram | 0 selected articles out of 0 |

| | |
|---|---|
| EEG | 0 selected articles out of 2 |

**KCE: Belgian health Knowledge Center**

All reports - KCE (fgov.be)

| | |
|---|---|
| Electroencephalography | 0 selected articles out of 1 |
| Electroencephalogram | 0 selected articles out of 0 |
| EEG | 0 selected articles out of 1 |
| électroencéphalographie | 0 selected article out of 1 |

**CEDIT, the Hospital-Based HTA Agency Of AP-HP**

Recommendations and Reports | Cedit (aphp.fr)

| | |
|---|---|
| Electroencephalography | 0 selected articles out of 0 |
| Electroencephalogram | 0 selected articles out of 0 |
| EEG | 0 selected articles out of 1 |
| électroencéphalographie | 0 selected article out of 0 |

**Haute Autorité de Santé**

**Haute Autorité de Santé - Résultat de recherche (has-sante.fr)**

| | |
|---|---|
| EEG | 1 selected article out of 218 |
| électroencéphalographie | 0 selected article out of 27 |

**Health Information and Quality Autority**

**Health Technology Assessments | HIQA**

| | |
|---|---|
| Electroencephalography | 0 selected articles out of 0 |
| Electroencephalogram | 0 selected articles out of 0 |
| EEG | 0 selected articles out of 0 |

**Irish Health Repository**

Lenus the Irish Health Repository

| | |
|---|---|
| Title: Electroencephalography AND epilepsy | 1 selected article out of 51 |
| Electroencephalogram | 0 selected articles out of 3 |
| Title: EEG AND epilepsy | 0 selected articles out of 51 |

**Norwegian Institute of Public Health**

**Norwegian Institute of Public Health - NIPH (fhi.no)**

| | |
|---|---|
| Electroencephalography | 0 selected articles out of 0 |
| Electroencephalogram | 0 selected articles out of 0 |
| EEG | 0 selected articles out of 3 |

**Swedish Agency for Health Technology Assessment And Assessment Of Social Services Home (sbu.se)**

| | |
|---|---|
| Electroencephalography | 0 selected articles out of 2 |
| Electroencephalogram | 0 selected articles out of 2 |
| EEG | 0 selected articles out of 4 |

**Healthcare Improvement Scotland**

**Healthcare Improvement Scotland**

| | |
|---|---|
| Electroencephalography | 0 selected articles out of 0 |
| Electroencephalogram | 0 selected articles out of 0 |
| EEG | 0 selected articles out of 0 |

**National Institute for Health and Care Excellence**

**NICE | The National Institute for Health and Care Excellence**

| | |
|---|---|
| electroencephalography AND epilepsy | 0 selected articles out of 2 |
| Electroencephalogram AND epilepsy | 1 selected article out of 5 |
| EEG | 0 selected articles out of 9 |

**NIHR Innovation Observatory**

**Innovation Observatory | Next generation search tools for the next generation. (nihr.ac.uk)**

| | |
|---|---|
| Electroencephalography | 1 selected article out of 2 |
| Electroencephalogram | 0 selected articles out of 1 |
| EEG | 0 selected articles out of 5 |

**National institute for health Research**

**Research Programmes (nihr.ac.uk)**

| | |
|---|---|
| electroencephalography AND epilepsy | 1 selected article out of 67 |
| Electroencephalogram AND epilepsy | 0 selected articles out of 67 |
| EEG | 0 selected articles out of 67 |

**Agency for Healthcare Research and Quality : Technology Assessment Program**

**Technology Assessment Program | Agency for Healthcare Research and Quality (ahrq.gov)**

| | |
|---|---|
| Electroencephalography AND epilepsy AND diagnosis | 0 selected articles out of 1 |
| Electroencephalogram AND epilepsy AND diagnosis | 0 selected articles out of 78 |
| EEG AND epilepsy AND diagnosis | 0 selected articles out of 83 |

**Agency for Healthcare Research and Quality : Evidence-Based Reports**

**Search Evidence-Based Reports | Agency for Healthcare Research and Quality (ahrq.gov)**

| | |
|---|---|
| Electroencephalography | 0 selected articles out of 0 |
| Electroencephalogram | 0 selected articles out of 0 |
| EEG AND epilepsy | 0 selected articles out of 4 |

**Google**

| | |
|---|---|
| intitle: Electroencephalography AND epilepsy AND machine learning AND diagnosis | 3 selected articles out of 9 |
| intitle: Electroencephalogram AND epilepsy AND machine learning AND diagnosis | 0 selected articles out of 9 |
| intitle: EEG AND epilepsy AND machine learning AND diagnosis | 1 selected articles out of 9 |
| intitle: Electroencephalography AND epilepsy AND algorithm AND diagnosis | 0 selected articles out of 9 |
| intitle: Electroencephalogram AND epilepsy AND algorithm AND diagnosis | 0 selected articles out of 9 |
| intitle: EEG AND epilepsy AND algorithm AND diagnosis | 0 selected articles out of 9 |
